# Performance of protein panels is inflated across many biomarker studies

**DOI:** 10.64898/2026.09.02.26362037

**Authors:** Lijun An, Caitlin A. Finney, Artur Shvetcov, the Global Neurodegeneration Proteomics Consortium (GNPC), Jacob W. Vogel

## Abstract

Data leakage is a prevalent yet underappreciated flaw in biomarker discovery studies. Through simulation and real-world proteomic data, we demonstrate that typical pipelines are broadly susceptible to this issue, producing inflated performance estimates, poor generalization, and excess false positives. We further introduce two tools to detect data leakage at the code and manuscript level, providing a practical path toward more rigorous and reproducible biomarker reporting.

## Main

The rapid growth of dementia populations worldwide urges scalable and accessible biomarkers for neurodegenerative diseases^1^. The success of phospho-tau markers for Alzheimer’s disease (AD)^2^ showcases how transformative such biomarkers are for scalable diagnosis and monitoring. However, sensitive fluid biomarkers remain under development for most neurodegenerative diseases. Recent advances in proteomic technologies have begun to address this gap by measuring thousands of potential biomarkers at once, yielding promising candidates such as secreted modular calcium-binding protein 1 (SMOC1) for early Alzheimer’s Disease^3^, YWHAG:NPTX2 synapse protein ratio for incipient cognitive decline^4^, and DOPA decarboxylase (DDC) for Lewy body diseases^5^. While many of these findings stem from sequencing cerebrospinal fluid (CSF), blood plasma offers a more promising opportunity for sequencing much larger and more diverse populations. Encouragingly, large-scale initiatives such as the Global Neurodegeneration Proteomics Consortium (GNPC)^6^ and UK Biobank^7^ are now creating an unprecedented opportunity for biomarker discovery, with the ultimate goal of identifying markers tracking disease status (e.g., diagnosis, progression, subtype) with high sensitivity, specificity, and generalizability.

The rapid expansion of biomarker discovery studies has not necessarily yielded a proportional uptick in clinically valid or clinically useful biomarkers, a concern increasingly recognized across the field^8^. Beyond the inherent biological complexity of identifying sensitive and reliable biomarkers, weaknesses in study design can further amplify this translational gap. These weaknesses include lack of external validation, selective reporting, and a critical analytical flaw known as “data leakage”.

Data leakage is unfortunately an inherent property of what has become the typical biomarker discovery pipeline (Fig. 1A). This pipeline comprises two main stages: 1) identification of candidate biomarker panels; and 2) evaluation of the resulting panel’s discriminative performance using machine learning (ML). Step 1 is normally achieved using mass univariate comparisons (e.g., differential abundance analysis[DAA]) across the full cohort, identifying significant biomarkers. These same significant markers are subsequently used for Step 2: training and validating ML models. These ML pipelines typically use train–test split or cross-validation frameworks within the same cohort, which is supposed to give an estimate of the panel’s performance in making predictions on unseen data. The problem with this typical pipeline is that the data is not actually “unseen”. When feature selection (Step 1) is performed on the full cohort before it is partitioned for cross-validation, information from future test samples leak into the model during training. Put simply, the analysis is circular; biomarkers are first selected because they differ between cases and controls, and the ML model is then tested on how well those same biomarkers distinguish cases from controls in the same data. In theory, this leads to inflation of performance estimates. Despite being a foundational premise in machine learning, data leakage nonetheless remains prevalent across many recent proteomics biomarker studies of aging and neurodegeneration published in high impact journals.

**Figure 1.**
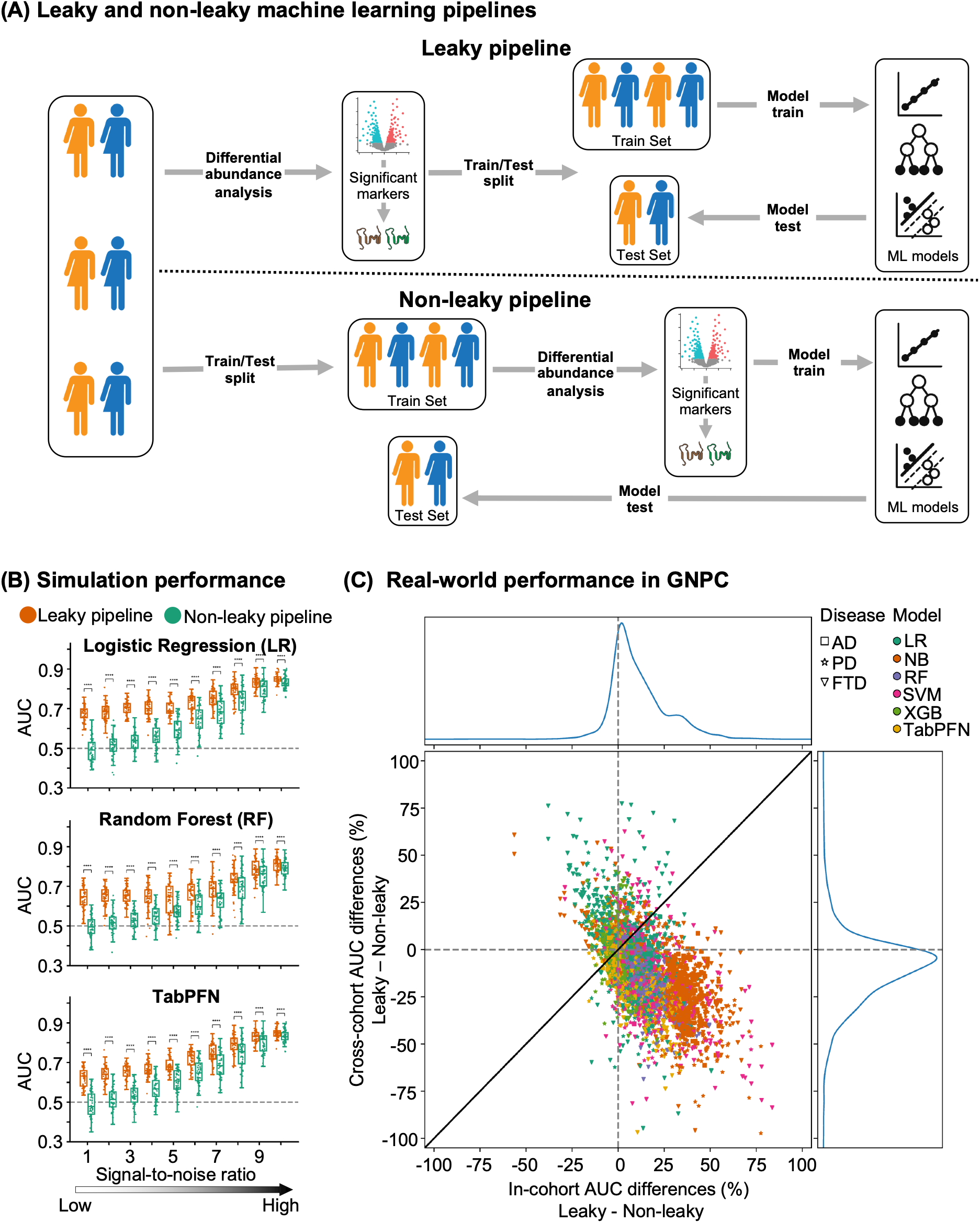
Leakage in most machine learning proteomics pipelines leads to systematic performance overestimation and worse generalizations. (A) Overview of the leaky and non-leaky analysis pipelines. Leaky pipeline: differential abundance analysis (DAA) is performed on the full cohort to identify “significant” markers, and the same cohort is then split into training and test sets for machine learning model development. Because the test-set samples contributed to marker selection, information from the test set leaks into the modeling process. Non-leaky pipeline: the cohort is first split into training and test sets to prevent leakage; marker selection and model development are performed exclusively within the training set, and the held-out test set is reserved only for final model evaluation. **(B)** Performance of three machine learning models on simulated case–control proteomics datasets across increasing signal-to-noise ratios. Orange points indicate a leaky pipeline and teal points indicate a non-leaky pipeline, in which feature selection was restricted to the training data. The x axis shows signal-to-noise ratio and the y axis shows area under the receiver operating characteristic curve (AUC); the dashed line marks chance performance (AUC = 0.5). Asterisks denote nominal significance from paired two-sided t-tests performed at each signal-to-noise level, without adjustment for multiple comparisons (****p < 0.0001). **(C)** Comparison of leaky and non-leaky pipelines in real-world plasma proteomics data from GNPC. Each point represents one model–cohort combination; shape denotes disease (AD, PD, FTD) and color denotes classifier (Logistic Regression: LR, NB: Naive Bayes, Random Forest: RF, SVM: Support Vector Machine, XGB, TabPFN). The x axis shows the percentage difference in AUC between leaky and non-leaky pipelines in within-cohort evaluation; values >0 indicate inflated performance under leakage. The y axis shows the corresponding percentage difference in AUC in across-cohort evaluation; values <0 indicate worse external generalization under leakage. The diagonal line indicates equality between within-cohort and across-cohort differences. Marginal density plots are shown for each axis.

Data leakage is well accepted as a methodological error among AI scientists, but the consequences of data leakage on reported biomarker performance and generalizability (and subsequent clinical translation) remains largely unquantified. Here, we ask the question: does leakage actually affect biomarker discovery pipeline in a meaningful way, or is it more of an inconsequential technological detail? To address this question, we systematically quantified the outcomes of data leakage in both simulated proteomics datasets (N = 500 simulations) and in a real-world multi-site proteomics datasets (GNPC; N = 7,548 participants spanning AD, PD, and frontotemporal dementia [FTD], across five separate clinical samples) using the Alzheimer’s Disease Data Initiative’s AD Workbench^9^. We benchmarked six widely used machine learning classifiers. Our goals were threefold – i) to establish whether data leakage actually inflates performance in real data; ii) to assess whether leakage affects a model’s ability to generalize to new sites or samples; and finally, iii) evaluate how well models with leakage perform in situations where greater-than-chance prediction should be impossible.

To evaluate the effects of data leakage under both controlled and real-world conditions, we first generated 50 proteomics datasets at each of ten signal-to-noise ratios (SNRs; 1–10). Each dataset included 100 disease and 100 control samples measured across 100 proteins. Five proteins were assigned true disease signals, with increasing separation between cases and controls as the SNR increased, whereas the remaining 95 proteins represented background noise to simulate sparse signal structure characteristic of real proteomics data. At SNR = 1, no true signal is detectable, whereas at SNR = 10, the five signal proteins achieve approximately 80% discriminative accuracy between disease and control groups. We then complemented this analysis using the real-world GNPC data across five sites and three neurodegenerative diseases. In this case, each disease-site combination was treated as an independent dataset. Within-site performance was assessed using five-fold cross-validation, whereas cross-site generalization was evaluated by training models in one site and testing them in the remaining sites. In both the simulated analysis and the GNPC analyses, we compared leaky and non-leaky pipelines (Fig. 1A). In the leaky pipeline, DAA was performed on the full dataset before cross-validation. The selected proteins were then used to train models on four folds and evaluate them on the held-out fold. In the non-leaky workflow, cross-validation splits were performed first. DAA and model training were then performed only within the training folds, while the held-out fold was used solely for evaluation. All models used default hyperparameters, and performance was summarized as the mean area under the receiver operating characteristic curve (AUC) across folds.

We explored these questions first using simulations where we could control all facets of the experiment, including the “actual” signal strength in the data (i.e. the upper limit on mean predictive performance). In our simulated experiments (Figure 1B), leaky pipelines consistently yielded higher classification accuracy than non-leaky pipelines across all classifiers and all signal strengths. This was expected behavior, since the leaky models had already been given information about the test set. A similar pattern emerged from our experiments on real-world GNPC data. From these experiments, we found that leakage inflated within-cohort performance by an average of 11 percentage points, with many instances exceeding 25 percentage points (x axis in Figure 1C). This degree of inflation is large enough to transform a biomarker panel from a modest performer to a clinically “promising” candidate, directly misleading research focus and translational prioritization.

Perhaps the most serious flaw introduced by leakage is the emergence of inadvertent false-positive biomarker panels in these settings. When no true biological signal was present in the simulated data (signal-to-noise ratio = 1; Figure 1B), correctly implemented pipelines performed no better than chance. Leaky pipelines, however, consistently produced “meaningful” classifications, with some runs achieving AUCs approaching 0.8, accuracies that would ordinarily be considered publishable. Crucially, these false positives arose consistently across all machine learning models, indicating that the problem is an intrinsic defect of the pipeline design rather than a particular modeling choice.

We finally investigated whether leakage leads to poorer generalization in unseen cohort validation, a rigorous test of clinical utility. When models trained under leaky pipelines were applied to unseen cohorts, performance dropped by an average of 10 percentage points (Figure 1C). This contrasts sharply with the 11 percentage point overestimation observed internally. Such divergence points to a worrying consequence of data leakage: it favours models that appear stronger on paper but are more likely to fail in the real world, ultimately hindering the identification of proteomic markers that generalize well in clinical practice. It is also important to note here that these experiments represent only the most basic examples of leakage; some publications contain far more egregious and impactful examples of leakage in their design.

Lack of replication and overoptimistic performance are a prevalent, if underdiscussed, concern in clinical biology and bioinformatics. We identify one common error, data leakage, that consistently explains part of this phenomenon. These findings raise several concerns with broad implications for the field of biomarker development. In the short term, inflated performance increases risk of misdirecting follow-up studies, funding decisions, and pharmaceutical investment towards candidates that will ultimately fail to replicate. In the long term, an accumulation of such errors could erode institutional confidence in proteomics as a reliable path to clinical discovery, and can affect public and industry confidence in academic reports of biomarker performance. More concerningly, our simulations show that leakage can generate false positive results even when no true biological signal exists. This is especially problematic for early-stage biomarker discovery, where effect sizes are often modest and independent validation is not always available. Finally, our real-world analyses indicate that biomarker panels developed using leakage pipelines generalize poorly across cohorts, impairing the translation from biomarker discovery to clinical deployment.

Why does leakage happen and why is it so common? One contributing factor is the considerable scale of modern proteomics data. Conventional biomarker studies focus on a small number of markers selected on the basis of prior biological knowledge, and are therefore informative by design. High-throughput proteomics, by contrast, is largely data-driven and identifying a robust panel from thousands of candidate proteins is a fundamental statistical challenge. Another factor is the methodological gap that emerges when proteomics studies increasingly rely on machine learning without sufficient cross-disciplinary expertise. As we mentioned earlier, avoiding data leakage is a foundational principle in machine learning, yet one that has been systematically overlooked in proteomics research. This concern is likely to grow as user-friendly computational packages and AI-assisted coding lower the barrier to complex analyses.

Addressing these issues requires coordinated action at both the study and publication stages. During study design and analysis, test sets should be strictly untouched until final evaluation. All feature selection, preprocessing and model tuning should be performed within the training data only, using nested designs where explicitly justified and necessary. Where possible, performance should also be evaluated in independent external cohorts, because cross-cohort validation provides a more realistic estimate of clinical robustness than internal resampling alone. Even in our own work^10^ where we rigorously avoided data leakage, we found that cross-validation performance within our sample was still optimistic compared to out-of-sample performance.

Beyond these design principles, leakage diagnostics should be incorporated into routine model development and reporting. We therefore developed *Leakly* (https://pypi.org/project/Leakly/), a lightweight Python package designed to detect data leakage in generic machine learning pipelines. *Leakly* operates by randomly permuting the outcome labels, thereby destroying any true association between the proteomics features and the prediction target. It then tests any user-specified machine learning pipeline or workflow to evaluate performance under these null conditions. Under permutation, a leak-free pipeline should perform no better than chance, since the labels carry no real signal. If a pipeline instead yields above-chance performance on permuted labels, this indicates that information from test samples has entered the training process. In this way, the model has inadvertently memorized spurious associations between features and the shuffled labels, a hallmark signature of leakage. *Leakly* provides a principled diagnostic that can be embedded directly into existing workflows without modification of the pipeline. Importantly, *Leakly* is robust to any kind of leakage, not just the kind exemplified here.

At the publication stage, manuscripts that make predictions from high-dimensional omics data should be assessed by reviewers with appropriate statistical or machine learning expertise, with explicit attention to data partitioning, feature-selection strategy, model tuning and external validation. These issues are often treated as technical details, but they can determine whether a reported biomarker signature is reliable or not. To further ease the burden on reviewers at the publication stage, we developed *Plumber* (https://demonlab-biofinder.github.io/Plumber/), a structured prompt generator that harnesses large language models (LLMs) to generate formatted methodological review reports.

The framework encodes a curated set of leakage indicators, including the feature-selection leakage described above, as pre-defined instructions. The LLM then applies these indicators systematically to manuscript methods. The output is a formatted report flagging either clear evidence of leakage or methodological ambiguities, providing reviewers and editors with a first-pass assessment before expert human review. This is particularly valuable given the well-recognised shortage of reviewers with machine learning expertise across biomedical fields. Although *Plumber* remains a prototype, we think that automated data leakage screening should become a first-round component of any journal’s editorial workflow for prediction studies.

Finally, open science should be viewed as part of methodological rigor rather than as an optional addition. Sharing code, models and, where ethically and legally possible, data or synthetic examples would allow independent verification, improve transparency and increase the long-term translational impacts of published studies. Finally, closer collaboration between clinicians, proteomics researchers, statisticians and machine learning scientists is essential. Many leakage problems are conceptually simple once recognized, yet they continue to appear because methodological designs are often inherited across fields without sufficient scrutiny. Building cross-disciplinary review and collaboration into biomarker discovery pipelines would help ensure that promising proteomic signatures are not only statistically impressive, but also reproducible, generalizable and clinically meaningful.

Although our analyses focus on proteomics and neurodegenerative disease, the implications extend across the biomedical sciences. Data leakage is not a proteomics-specific problem. Any high-dimensional omics discipline, such as genomics, transcriptomics, metabolomics or radiomics that combines large feature spaces with machine learning classifiers, faces the same structural vulnerability. As multi-modal data integration and AI-assisted analysis become popular practice across clinical research, the risk of systematically inflated, non-replicable findings grows proportionally. The biomedical community has invested heavily in open data, pre-registration, and reproducibility initiatives following the replication crisis in psychology^11^ – it would be a costly mistake to allow the same crisis to take root in molecular medicine. The fixes are neither technically demanding nor resource-intensive. We only need principled pipeline design, cross-disciplinary literacy, and a shared commitment to methodological rigor as a non-negotiable standard. Getting the statistics analysis right is not a barrier to discovery but rather the foundation on which robust clinical discoveries are built.

## Data Availability

GNPC (https://www.neuroproteome.org/) is open access.

## Acknowledgment

This work was supported by the SciLifeLab and Wallenberg Data Driven Life Science Program (grant no. KAW 2020.0239 to J.W.V.) the Swedish Research Council (grant no. 2024-03642 to J.W.V.), and the Biwas Family Foundation (L.A. and J.W.V.). Data discovery and/or analysis services contributing to this work were provided in-kind by the AD Data Initiative. Our computational work was supported by project sens2023026 by the National Academic Infrastructure for Supercomputing in Sweden (NAISS; https://www.naiss.se) at UPPMAX, project NAISS 2025/23-728 and NAISS 2025/22-1279 by Chalmers e-Commons at Chalmers and the Berzelius resource (Berzelius-2026-231) funded by the Knut and Alice Wallenberg Foundation at the National Supercomputer Centre. NAISS is partially funded by the Swedish Research Council through grant agreement no. 2022-06725. The funding sources had no role in the design and conduct of the study; in the collection, analysis and interpretation of the data; or in the preparation, review or approval of the paper.

